# Whole-genome sequencing of SARS-CoV-2 in the Republic of Ireland during waves 1 and 2 of the pandemic

**DOI:** 10.1101/2021.02.09.21251402

**Authors:** P.W.G. Mallon, F. Crispie, G. Gonzalez, W. Tinago, A.A. Garcia Leon, M. McCabe, E. de Barra, O. Yousif, J.S. Lambert, C.J. Walsh, J.G. Kenny, E. Feeney, M. Carr, P. Doran, P.D. Cotter, on behalf of the All Ireland Infectious Diseases Cohort Study and the Irish Coronavirus Sequencing Consortium.

**Affiliations:** Centre for Experimental Pathogen Host Research, School of Medicine, University College Dublin, Ireland; St Vincent’s University Hospital, Dublin, Ireland; Teagasc Food Research Centre, Moorepark, and APC Microbiome Ireland, Cork, Ireland; National Virus Reference Laboratory, University College Dublin, Ireland; International Collaboration Unit, Research Center for Zoonosis Control, Hokkaido University, Sapporo, Japan; Teagasc Grange, Animal and BioScience Department, County Meath, Ireland; Dept of International Health & Tropical Medicine, Royal College of Surgeons in Ireland, Dublin, Ireland; Beaumont Hospital, Dublin, Ireland; Wexford General Hospital, Wexford, Ireland; Mater Misericordiae University Hospital, Dublin, Ireland; Clinical Research Centre, School of Medicine, University College Dublin, Ireland

**Keywords:** SARS-CoV-2, COVID-19, whole genome sequencing, sequencing, variant, lineage

## Abstract

**Background:** Whole-genome sequencing (WGS) of SARS-CoV-2 laboratory-confirmed cases can provide insights into viral transmission and genetic diversity at a population level. However, less is known about the impact of non-pharmaceutical interventions (NPIs), including ‘lockdowns’, on circulating SARS-CoV-2 lineages and variants of concern, the relative contribution of travel to re-emergence of pandemic waves within communities or how different lineages and variants contribute to disease severity.

**Methods:** We have conducted an analysis within a prospective, multicentre observational study of individuals attending four hospitals in the South-East of Ireland with COVID-19. Samples underwent WGS from which lineages and variants were assigned, lineage frequency was plotted over time and phylogenetic analysis was employed to determine the origin of variants detected post-lockdown. Univariate and multivariate analyses assessed relationships between viral lineage/variant and COVID-19 disease severity.

**Results:** We analysed 225 genome sequences across two SARS-CoV-2 waves, 134 (59.6%) from wave 1 (March to June) and 91 (40.4%) from wave 2 (July to December), representing 15.2% of COVID-19 admissions to these hospitals during the sampling periods. Four variants (B.1.1.162, B1.1.70, B.1.1.267 and B.1.1) comprised 68% of variants detected during wave 1. Of these variants, only a single B.1.1.70 sequence was detected in wave 2, while the B.1.177 lineage emerged and contributed to 82.3% of lineages detected. Phylogenetic analysis suggested multiple introductions of wave 2 variants from outside Ireland. We found no consistent association between SARS-CoV-2 lineages and disease severity.

**Conclusions:** These data suggest elimination of common SARS-CoV-2 lineages from hospitalised cases associated with effective NPIs and that importation of new viral variants through travel was a significant contributor to the re-emergence of the pandemic in the second wave in Ireland. Our findings highlight the importance of genomic surveillance in identifying circulating viral genetic diversity and variants of concern and, also, modelling the disease burden of SARS-CoV-2.

## Introduction

More than a year after being first reported in Wuhan, Hubei province, China, [1] the COVID-19 pandemic, caused by severe acute respiratory syndrome coronavirus 2 (SARS-CoV-2) continues to cause significant morbidity and mortality worldwide. In the Republic of Ireland, the first case of community transmission of SARS-CoV-2 was identified on 21st February 2020 [2]. Since then, at the time of writing (Feb 7, 2021), over 201,763 cases have been reported, linked to 3,621 deaths (www.hpsc.ie).

The pandemic in Ireland has been characterised by three distinct ‘waves’ of infections, each controlled by non-pharmaceutical interventions (NPIs) involving a series of societal restrictions comprising physical distancing, hand hygiene and limiting movements within the country. During the first wave, implementation of these restrictions (or ‘lockdowns’) was associated with significant reductions in daily reported cases and mortality, followed by phased relaxation in restrictions [3]. Despite easing of restrictions on 18th May, this impact was maintained, with lowest daily reported case numbers of 3 laboratory-confirmed cases reported at the end of June 2020 [4]. This was achieved while permitting international travel through the country’s airports and seaports and permitting travel across the land border between the Republic of Ireland and Northern Ireland. Following phase three easing of restrictions in late June, which included permission to travel within Ireland, case numbers began to rise and subsequent efforts at control of transmission through community restrictions have failed to achieve the effects seen in June 2020. The highest (level 5) restrictions were implemented on 21st October 2020 with the lowest subsequent reported case numbers at the end of ‘wave 2’ dropping to 183 on 3rd December 2020, at the time when level 5 restrictions began to be relaxed.

Since first being reported in China, the evolution of the virus through mutations has enabled epidemiological mapping of the virus through phylogenetic analyses of viral genome sequences. This genomic epidemiology provides invaluable insights into the origins of viral transmissions within countries, and the impact of the mutations on either transmission or disease severity. For example, the identification of the 20AEU1 or B.1.177 variant, first identified in Spain on June 20^th^, 2020, was subsequently reported across Europe [5]. Although not linked to increased transmission or mortality, its increased prominence in the subsequent months across Europe supported holiday-related travel as an important factor in its spread. Since then, additional variants of concern, such as the B.1.1.7, B.1.351, and P.1 variants have been identified, [6, 7] which have been linked to both increased transmissibility and potentially worse clinical outcomes [8].

Although a number of studies have described genotypic variation in SARS-CoV-2 across Europe, few have been able to track the impact of effective NPIs, such as an effective lockdown as occurred during the first wave in Ireland, on viral genetic diversity in a representative sample of reported cases, and few studies have related differences in viral lineage to clinical outcomes [9]. Our objectives in the present study were to describe the genetic variation in SARS-CoV-2 lineages among individuals hospitalised with COVID-19 in the East and South-East of Ireland during 2020, covering two of the three ‘waves’ of the SARS-CoV-2 pandemic in Ireland, to define the impact of lockdown on lineage diversity and to explore the geographical source of infections during the course of the pandemic.

## Methods

We analysed samples taken from subjects enrolled in the All-Ireland Infectious Diseases (AIID) Cohort, a multicentre, prospective, longitudinal, observational cohort that enrols consecutive adult subjects (>18 years old) attending hospitals services for management of infections. The AIID Cohort recruits from eleven clinical centres across Ireland. Subjects provide consent for use of routine clinical and laboratory data for research, including demographic characteristics, symptom history, comorbidities, disease severity, clinical outcome, epidemiological risk and treatments, along with results of routine laboratory and radiological investigations. In addition, subjects consent to collection and biobanking of biological samples, including respiratory samples such as nasopharyngeal swabs. For this analysis, four participating hospitals located in Dublin and the South-East of Ireland provided samples; the Mater Misericordiae University Hospital located in central Dublin, St Vincent’s University Hospital located in south Dublin, Beaumont University Hospital located in north Dublin and Wexford General University Hospital in the south-east of Ireland. The AIID Cohort is approved by local institutional review boards and all participants provide written, informed consent. Data and samples within the AIID Cohort are accessed through standardised Data Access Guidelines and all approved Data Access Requests are approved by the local Ethics Committee.

The Irish Coronavirus Sequencing Consortium (ICSC) was established in early 2020 to facilitate large scale sequencing of SARS-CoV-2 samples to map the evolution of the virus in Ireland. The ICSC collaboration involves academic and commercial laboratories across the Republic of Ireland, with linked bioinformatic analysis. This analysis represents an approved AIID Access Request derived from a collaboration between the AIID Cohort and the ICSC.

From biobanked nasopharyngeal samples stored in viral transport medium taken from the AIID Cohort subjects with laboratory-confirmed SARS-CoV-2 infection (qRT-PCR positive cases) RNA was extracted using the automated platform for nucleic acid extraction EX3600 (Liferiver Biotech, Shanghai, China). Following RNA extraction, samples underwent qRT-PCR for SARS-CoV-2 using primers directed against the RNA-dependent RNA polymerase (RdRp) region of the viral genome on using the COVID-19 genesig® Real-Time PCR assay (Primerdesign Ltd, Hampshire, United Kingdom), on the The LightCycler® 480 PCR platform (Roche Diagnostics, Basel, Switzerland). Samples with sufficient RNA (defined as a positive SARS-CoV-2 PCR with cycle thresholds (Cq) ≤30 cycles) were forwarded for sequencing.

Following RNA extraction and qRT-PCR testing to ensure sufficient RNA for sequencing, single stranded cDNA was synthesised from samples with a Cq< 30 by reverse transcription with Superscript IV RT (Invitrogen, UK). This cDNA was used to generate sequencing libraries using a method based on the ARTIC amplicon sequencing protocol for COVID-19 [10, 11]. In brief, amplicons spanning the SARS-CoV-2 genome were amplified using ARTIC V3 primers. The amplicons were end-repaired using the Ultra II End prep enzyme (New England Biolabs (NEB), MA) and then native barcodes (Oxford Nanopore Technologies, Oxford, United Kingdom; ONT) were added by ligation using the NEBNext Ultra II Ligation Master Mix and Enhancer (NEB). Following barcoding, amplicons for each sample were pooled and cleaned using Ampure beads (Beckman Coulter, CA) and ONT sequencing adaptors added by ligation using the Quick T4 DNA ligase (NEB). The adaptor-ligated libraries were then cleaned using Ampure beads and pooled libraries loaded on flowcells and sequenced on either the MinION or GridION devices (ONT) for 24-72 hrs. Following sequencing, Fast5 files were uploaded to the Irish Centre for High End Computing (ICHEC) and bioinformatics analysis including genome assembly and variant calling were performed as described by the ARTIC network [12].

FastA files of SARS-CoV2 whole genome consensus sequences were used for assignment of a Pangolin lineage to each sample. The PANGOLIN nomenclature offers an algorithmic method to classify, distinguish and put in context the different SARS-CoV-2 sequences considering the large amount of available genomic sequence data [13] in public databases, such as GISAID (www.gisaid.org) [14]. However, as the characterisation of the different SARS-CoV-2 lineages become available and more sequences are accumulated retrospectively, some of these lineages are revised and reassigned [15]. Consequently, the following descriptions are subject to revisions to such a dynamic nomenclature. The lineages assigned in this report were based on the pangoLEARN version 2021/02/01. For reference, the characteristic non-synonymous mutations in the SARS-CoV-2 spike protein associated with these lineages are provided in Table 1. Lineages were assigned to samples, aligned with metadata from the AIID Cohort (age, gender and county of residence), and genome sequences were uploaded onto the public database GISAID.

**Table 1:** Characteristic amino acid mutations in the spike protein for PANGOLIN lineage assignment.

| PANGOLIN Lineage | GISAID Lineage | Amino acid mutations |
| --- | --- | --- |
| B.1.1 | G | D614 |
| B.1.1.267 | GR | D614 |
| B.1.177 | GV | D614, A222V |
| B.1.177.18 | GV | D614, A222V, E583D |
| B.1.1.7 | GR | D614, A570D, D1118H,<br>H69del, N501Y, P681H,<br>S982A, T716I, V70del,<br>Y145del |

### Phylogenetic analysis

Global genomic sequences were downloaded from GISAID [14]. Of these genomes, those with two or less nucleotide differences to any of the Irish B.1.177 sequences were selected. Subsequently, 310 sequences with the earliest sampling dates were chosen and multiple sequences aligned to the reference (MN908947) and 71 AIID B.1.177 genomic sequences with MAFFT [16]. A phylogenetic tree was inferred from this multiple sequence alignment with RAxML [17] with the general time reversible model of substitution allowing for heterogeneity among sites (GTRCAT).

### Statistical analysis

We used summary statistics as appropriate and bubble plots to describe the patterns of distribution of genome lineages over time (month or week from 24^th^ March to 27^th^ December 2020). Bubble charts were created in R with ggplot2 packages [18]. We used ordinal logistic regression (proportional odds model) to examine the associations between the most frequently identified lineages and a primary outcome measure of maximal COVID-19 disease severity defined according to the WHO guidance [19] and collapsed into mild, moderate and severe for analysis. The cumulative odds ratios (OR) for the proportional odds logistic regression represented the cumulative odds of a more severe clinical disease (“Severe vs Moderate/Mild disease, or “Severe/Moderate vs Mild disease). The proportional odds assumption were tested using the chi-square score test and graphical assessments to check for parallelism. All statistical analyses were performed in SAS version 9.4 (SAS Institute), and R version 4.0.2 (Vienna, Austria) [20].

## Results

Between 24^th^ March and 27th December 2020, 1,211 subjects were recruited to the AIID Cohort, of which 843 (69.6%) were laboratory-confirmed SARS-CoV-2 positive by qRT-PCR. Of these, 275 subjects from four participating hospitals provided 660 respiratory samples for biobanking of which 225 were suitable for genome sequencing. Demographics of participating subjects broadly reflected the demographics of hospitalised cases of COVID-19 in Ireland (Table 2), with an average age of 65 years, predominantly male (55%) and Caucasian (83%). Greater than 80% of subjects reported an underlying condition, with hypertension, chronic respiratory disease, chronic heart disease and malignancy being the most common underlying conditions reported. A substantial portion of subjects experienced either moderately severe or severe COVID-19 infection, with an overall 10.6% mortality rate.

**Table 2.**
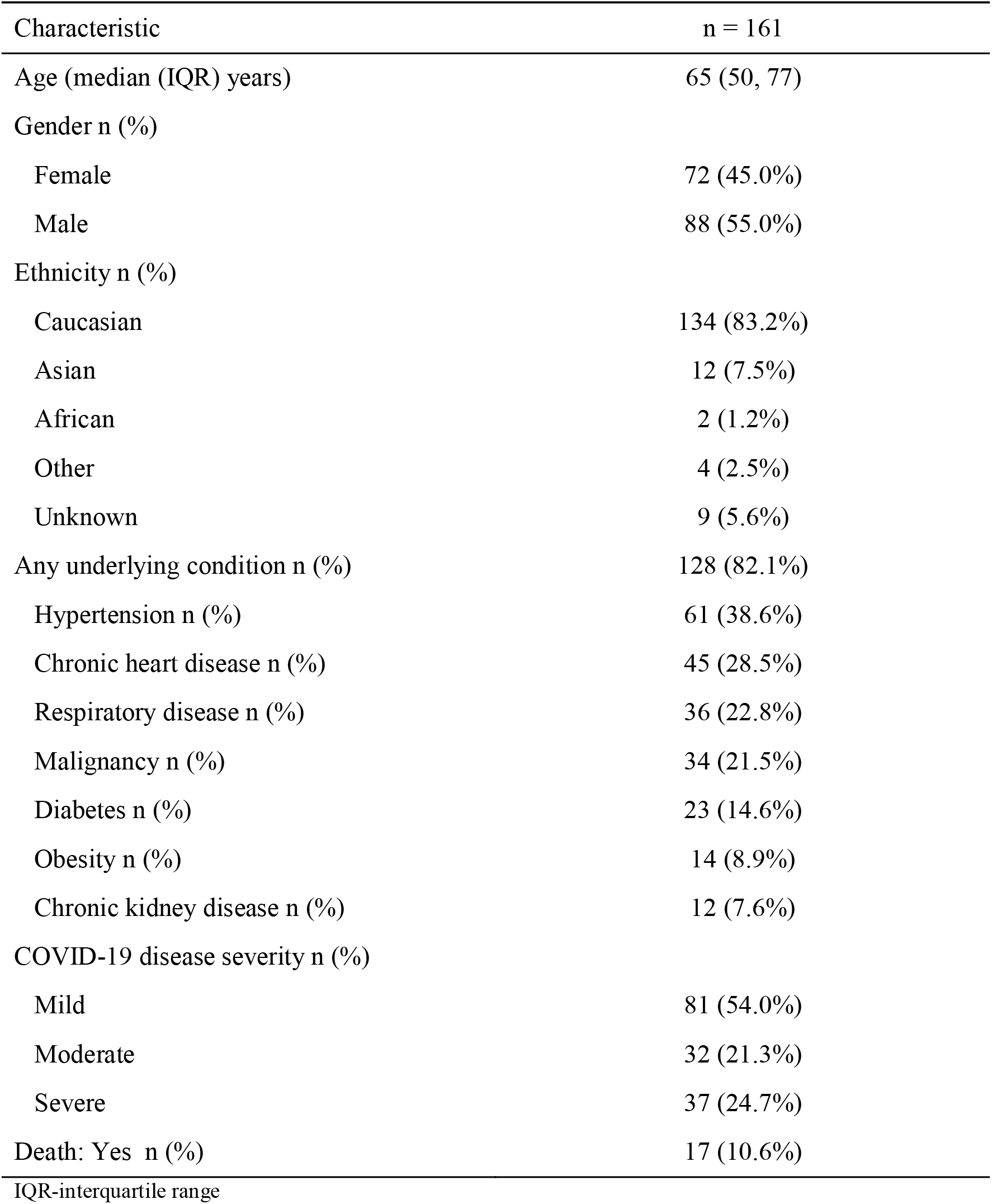
Characteristics of the study population

| Characteristic | n = 161 |
| --- | --- |
| Age (median (IQR) years) | 65 (50, 77) |
| Gender n (%) |  |
| Female | 72 (45.0%) |
| Male | 88 (55.0%) |
| Ethnicity n (%) |  |
| Caucasian | 134 (83.2%) |
| Asian | 12 (7.5%) |
| African | 2 (1.2%) |
| Other | 4 (2.5%) |
| Unknown | 9 (5.6%) |
| Any underlying condition n (%) | 128 (82.1%) |
| Hypertension n (%) | 61 (38.6%) |
| Chronic heart disease n (%) | 45 (28.5%) |
| Respiratory disease n (%) | 36 (22.8%) |
| Malignancy n (%) | 34 (21.5%) |
| Diabetes n (%) | 23 (14.6%) |
| Obesity n (%) | 14 (8.9%) |
| Chronic kidney disease n (%) | 12 (7.6%) |
| COVID-19 disease severity n (%) |  |
| Mild | 81 (54.0%) |
| Moderate | 32 (21.3%) |
| Severe | 37 (24.7%) |
| Death: Yes n (%) | 17 (10.6%) |
| IQR-interquartile range |  |

Sequenced samples represented subjects residing in 12 counties in Ireland, including Clare, Dublin, Galway, Kildare, Longford, Louth, Meath, Monaghan, Offaly, Waterford, Wexford and Wicklow, with Dublin (68%), Wicklow (21%) and Wexford (4%) contributing the greatest proportion of samples. Demographics of AIID Cohort subjects contributing sequenced samples are outlined in Table 1.

During the months where clinical sites provided samples for genome sequencing, the overall number and proportion of hospital admission that were sequenced was 6/37 (16.2%) at Wexford, 21/566 (3.7%) at MMUH, 137/450 (30.4%) at SVUH and 11/96 (11.5%) at the Beaumont hospital site. This represents an overall 15.2% of COVID-19 admissions where sequencing data is available.

Twenty-six lineages from 225 cases were identified between the period 24^th^ March and 27th December 2020 (Figure 1), with cases concentrated in Dublin, Wicklow and Wexford (Figure 2). Five lineages accounted for 81.8% of cases, these were B.1.177 (including the B.1.177.18 sublineage, 34.2%), B.1.1.267 (20.0%), B.1.1 (including sublineages B.1.1.70, B.1.1.162 and B.1.1.247, 22.2%), B (3.6%) and B.40 (3.6%).

**Figure 1:**
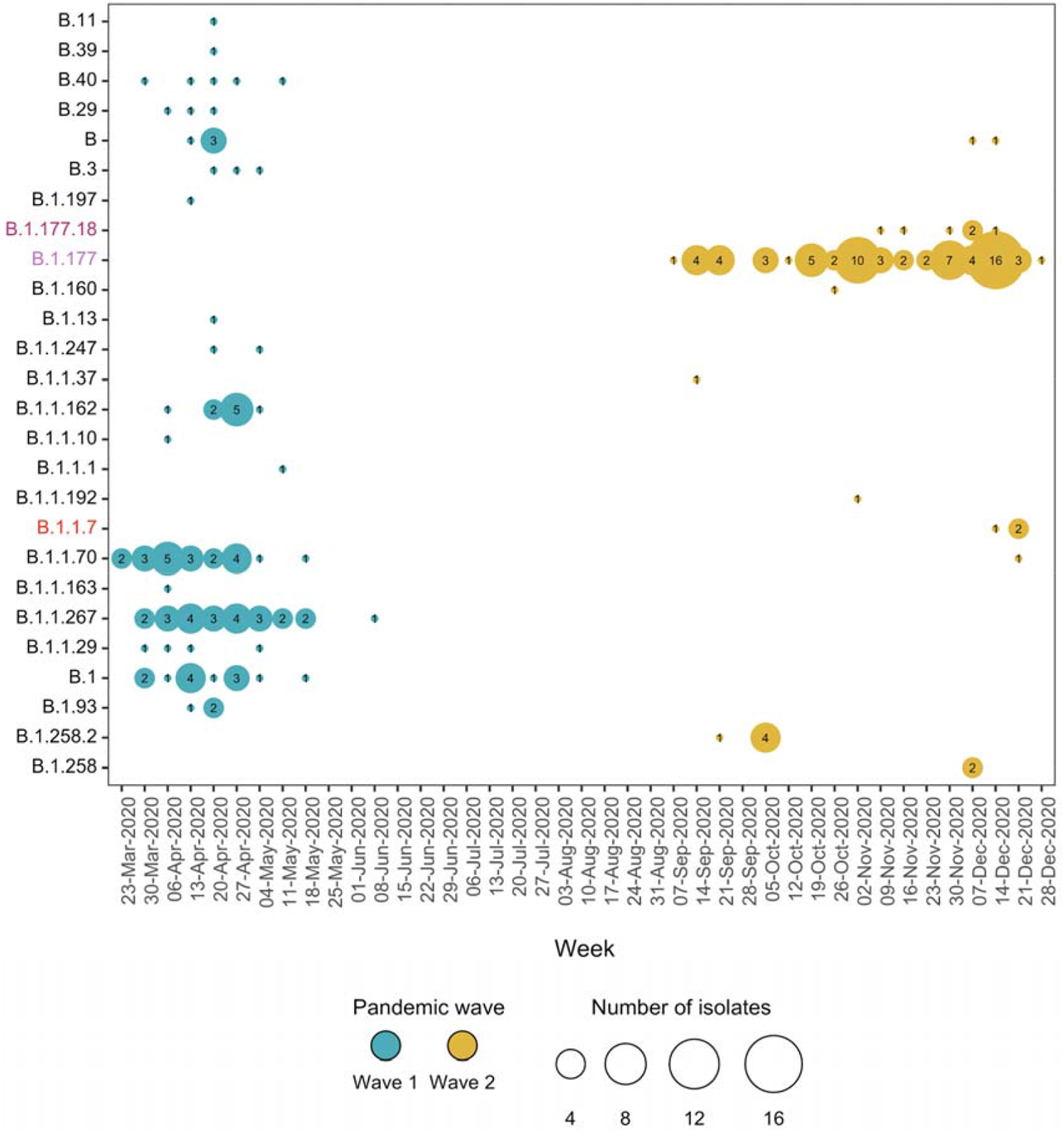
Pattern of SARS-CoV-2 genome lineage classification by week between March and December 2020 The size of the circle is proportional to the number of lineages detected.

**Figure 2:**
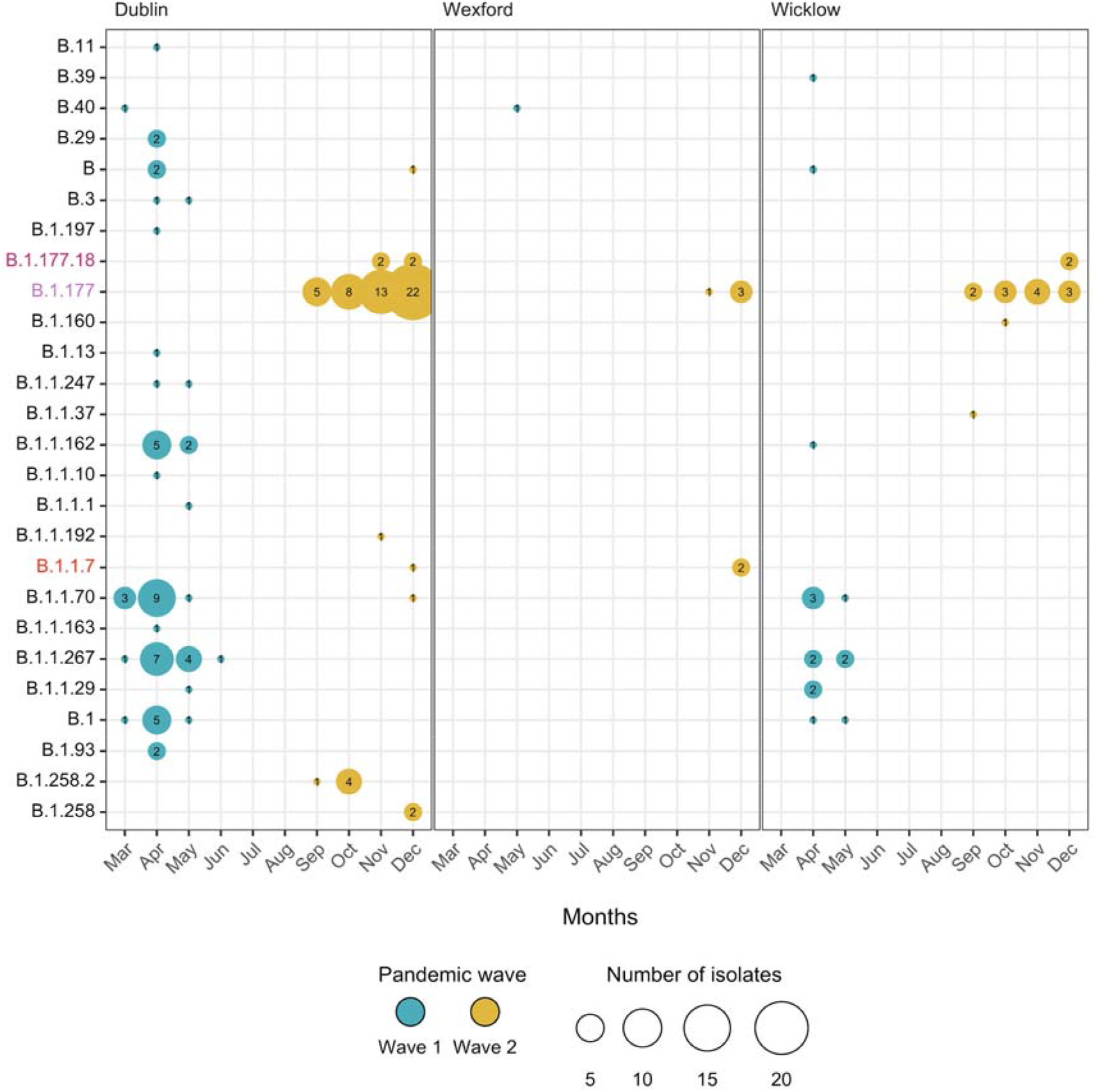
SARS-CoV-2 genome lineage classification by month and county of residence between March and December 2020 The size of the circle is proportional to the number of lineages detected.

**Figure 3:**
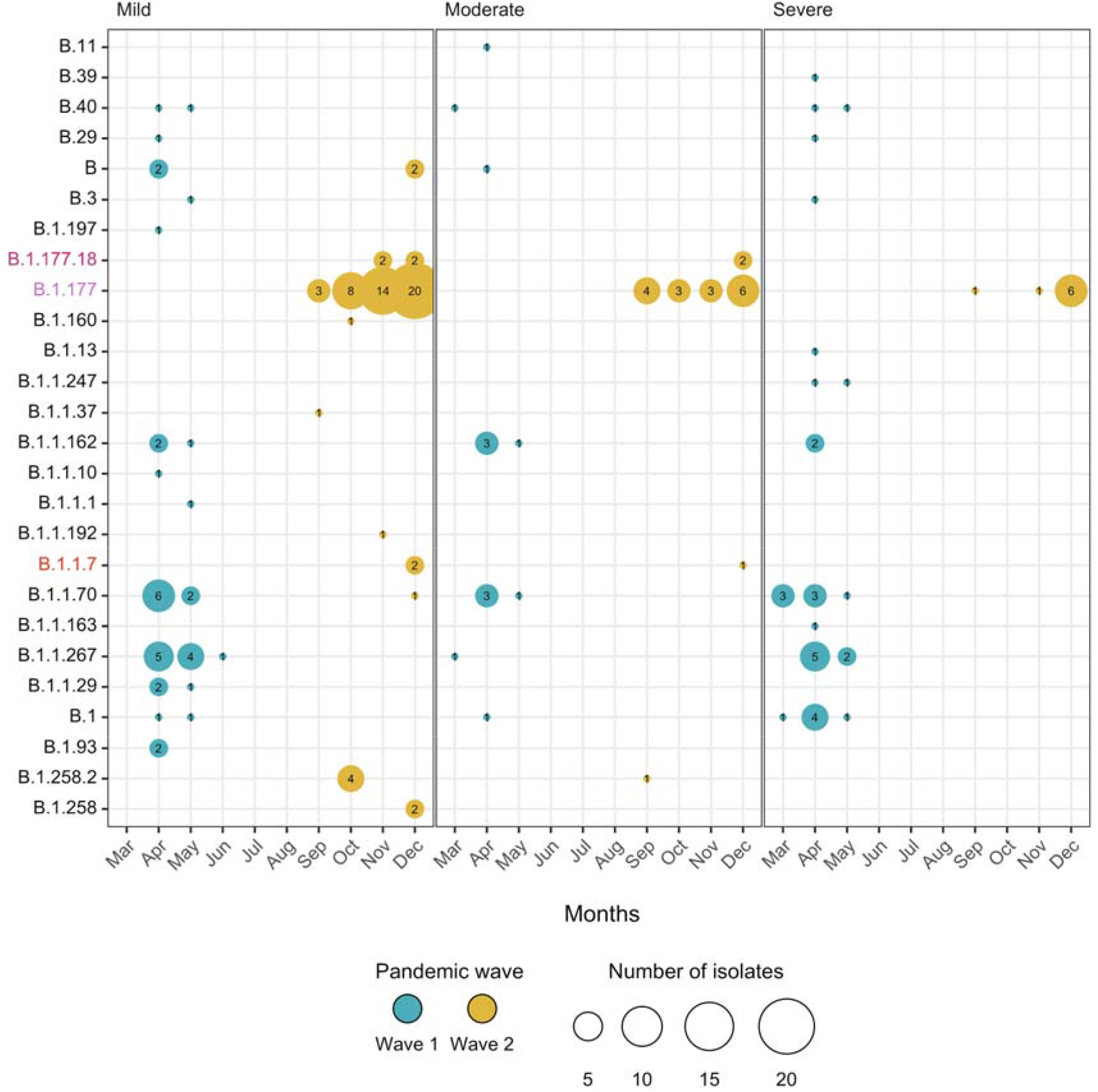
SARS-CoV-2 genome lineage classification by month and COVID-19 disease severity grade between March and December 2020 The size of the circle is proportional to the number of lineages detected.

Of the 131 cases analysed from the first SARS-CoV-2 wave, dating from March to the end of June 2020, two viral lineages represented 68.7% of sequences: B.1.1.267 (33.6%) and B.1.1 (37.4%). However, by mid-June 2020 these variants were no longer detected (B.1.1.267 last reported 6th June, B.1.1 (sublineage B1.1.162) on 8th May), coinciding with decline in reported cases nationally that accompanied the country-wide lockdown.

From July onwards, of 91 sequenced samples from the second wave, the predominant SARS-CoV-2 lineage reported was B.1.177 (including the B.1.177.18 sublineage), first reported September 10th, comprising 82.4% of lineages, followed by B.1.258.2 (5.5%) and B.1.1.7 (3.3%) (Figure 1). None of the common SARS-CoV-2 viral lineages detected during the first wave re-emerged during the second wave, with the exception of a single case of B.1.1.70 which did not arise until 21st December 2020.

Sequences from the first wave in lineages B.1.1.70 (n=20) and B.1.1.267 (n=14) have a within mean group distance of 4.9 nucleotide mutations. On the other hand, sequences from the second wave in lineages B.1.177 (n=66) and B.1.177.18 (n=5) evidenced a within mean group distance of 11.9 nucleotide mutations; such a disparity in the divergence accumulated in sequences of both waves is consistent with multiple introductions of B.1.177 members with some divergence among them.

Phylogenetic analysis of B.1.177 genomes from our analysis, when compared to genome sequences available in GISAID that had less than two mutations separating from our cases, supported multiple introductions of this lineage to Ireland from different European countries (Figure 4), in particular from the United Kingdom, including a small number of B.1.177.18 identified as English cases. In addition, monophyletic clusters of Irish samples also reflect some divergence accumulated through community transmission. Overall, these data suggest multiple sources of origin, consistent with multiple, distinct introductions of these variants into Ireland (Figure 4), likely through travel.

**Figure 4.**
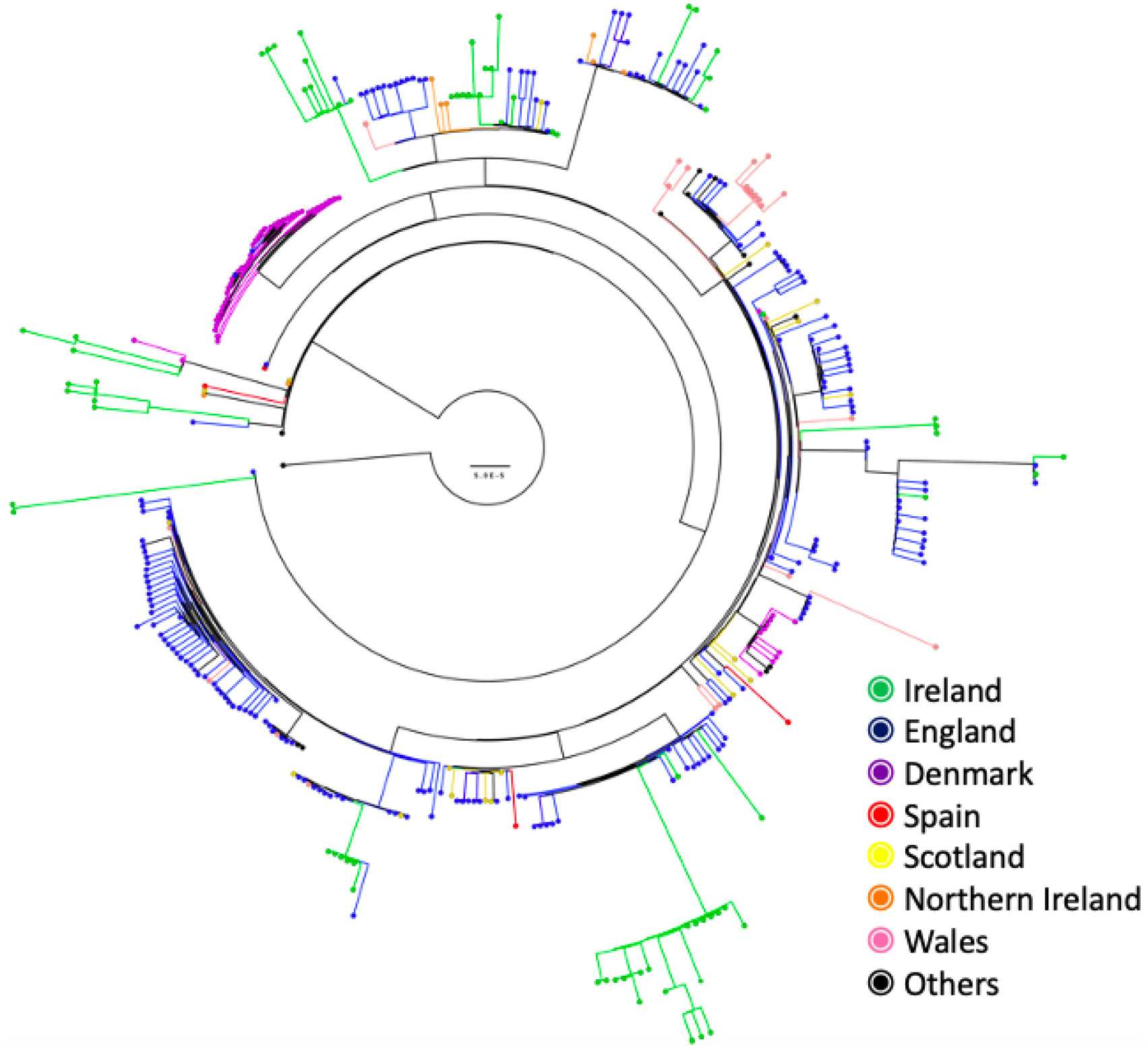
Circular phylogenetic tree of AIID Irish SARS-CoV-2 lineage B.1.177 genomic sequences and international closest sequences. The phylogenetic tree is rooted with the Wuhan reference genome (MN908947) and tips showing the country of origin are coloured green (Ireland), blue (England), purple (Denmark), red (Spain), yellow (Scotland), orange (Northern Ireland), pink (Wales) and black (others), respectively. The scale bar represents 5.0E-5 nucleotide mutations per site.

Although the B.1.1.7 variant of concern, first reported in Kent in the UK from samples taken in late September 2020 [21] was detected in only 3 (1.3%) samples sequenced, all were reported in the 10 days leading up to 27th December 2020 and comprised 15% of samples sequenced between those dates. The B.1.1.7 variant with the spike N501Y amino acid substitution was first identified in our sample set on 17th December 2020 from a subject with symptom onset dating back to the 10th December 2020.

In initial, unadjusted proportional odds models, compared to the rest of the sample, those with the B.1.1.267 lineage had a significantly increased odds of experiencing a more severe clinical disease (OR=3.24; 95% CI; 1.06-9.92), while those with the B.1.1 lineage had borderline significantly increased odds of experiencing more severe clinical disease (OR=2.01, (95% CI; 0.98-4.39). In contrast, those cases infected with the B.1.177 lineage, predominant during the second wave of infections, had significantly reduced odds (OR=0.49, 95%; 0.26-0.92) of experiencing a more severe clinical disease in comparison to those without the lineage. However, these strength of these associations were abrogated by correction for patient demographics (age, gender, ethnicity and presence of comorbidities), no longer being statistically significant after adjustment.

## Discussion

This is the first study to describe the change in lineages of SARS-CoV-2 in Ireland in 2020 from a representative sample of cases linked to clinically relevant data. Our analyses reveal three main findings; that, compared to the second wave, the first SARS-CoV-2 wave in Ireland was characterised by a larger number of distinct lineages, that the use of successful NPIs in the form of an effective ‘lockdown’ was accompanied by the disappearance of these lineages from those contributing to hospitalisations among the participating hospitals, and that the second wave was derived from viral genetic lineages that were most closely phylogenetically linked to genotypes outside of Ireland, suggesting multiple introductions through travel during the summer of 2020.

We observed distinct lineages of SARS-CoV-2 contributing to wave 1 and wave 2 of infections. The B.1.1 and B.1.1.267 lineages represented the majority of genome sequences identified from cases during wave 1. The B.1.1 lineage was first reported in Europe as early as January 24^th^ following the start of the Italian outbreak attributed largely to this lineage [22]. B.1.1 members possess the characteristic mutation A→G at nucleotide 23403 (based on the reference genome MN908947) resulting in an aspartic acid to glycine substitution at amino acid residue 614 of the spike protein (D614G), which has been observed as prevalent in nearly all samples sequenced in Ireland [23]. This mutation has been shown to increase viral fitness while maintaining similar morphology and *in vitro* neutralization properties [24, 25]. The B.1.1.267 lineage, genotypically similar to B.1.1 (also containing the D614G substitution), was the most common reported lineage during the first wave and has been identified solely in Ireland according to the PANGOLIN lineage website [15]. Similarly, members in the B.1.1.162 lineage were first reported in mid-February 2020 and denominated a United Kingdom lineage [15]. These data suggest that commonly reported lineages during wave 1 contributing to hospitalisations in our cohort included a mixture of originally imported lineages from continental Europe along with viruses that had evolved within Ireland.

With the introduction of effective lockdown in March 2020 in Ireland, the number of reported cases and associated hospitalisations dropped significantly, with the lowest reported daily case numbers (3 cases) recorded at the end of June, coinciding with a 14 day incidence rate falling to one of the lowest in Europe (2.77/100,000 in week 26 of 2020 [26]. The success of this lockdown in suppressing transmission was reflected in the drop off in hospitalisations of COVID-19 hospitalisations among our cohort.

However, as reported cases in Ireland began to increase from the end of June onwards, [26] samples from hospitalised patients in the cohort no longer derived from the sequences that comprised much of wave 1. In contrast, the B.1.177 lineage (and the B.1.177.18 sublineage) were detected predominantly in our hospitalised cohort, representing 82.4% samples analysed from the second wave. This lineage has been traced to Spain in early March; however, it became widespread across Europe during July and August 2020 [5]. Our phylogenetic analysis points to multiple introductions of this lineage into Ireland from overseas, in particular from the United Kingdom, with smaller clusters within Irish samples also reflecting continued divergence of this lineage through onward community transmission as it became established in Ireland.

Taken together, these data suggest that the first lockdown may have largely eradicated these commonly reported lineages from the circulating pool of transmitting viruses contributing to hospitalisations in the south-east Ireland, and that the second wave of infections resulting in hospitalisations were largely seeded by multiple, travel-related events from countries outside of the island of Ireland.

The importance of travel contributing to changes in lineages and variants of concern is further underlined by the detection of the B.1.1.7 variant in a cohort subject who first experienced symptoms in early December. The B.1.1.7 lineage was first sequenced in September in the United Kingdom; however, its spread and higher transmissibility were reported on December 20^th^ [27]. The lineage B.1.1.7, also known as SARS-CoV-2 VUI 202012/01 (Variant Under Investigation, year 2020, month 12, variant 01), has grown in frequency in the United Kingdom becoming the most prevalent lineage with a similar trend also observed in other European countries, including Ireland and Denmark [28]. Members of this lineage are characterised by a higher number of characteristic, non-synonymous, amino acid mutations in the spike protein (Table 1) that have been hypothesized to have originated from virus evolution in a chronically-infected individual [21].

Among the mutations described in the B.1.1.7 variant, the N501Y mutation, localised in the receptor binding site has been shown to increase the binding of SARS-CoV-2 to the angiotensin-converting enzyme 2 (ACE2) host receptor [29], inferring increased viral fitness and transmission properties. Although some data suggest that this variant may escape the neutralisation by select convalescent sera and therapeutic monoclonal antibodies, sera from participants of a trial of the mRNA based COVID vaccine BNT162b2 neutralised with similar efficacy the wild type N501 and the variant N501Y [30] suggesting retained vaccine efficacy against the B.1.1.7 variant.

We explored associations between presence of specific SARS-CoV-2 lineages and maximal disease severity experienced by cohort participants, according to WHO classification [19]. Data on relationships between SARS-CoV-2 lineages and disease severity are lacking; we could find no prior reports analysing relationships between B.1.1.269 and COVID-19 disease severity. Although univariate analyses did suggest that the B.1.177 lineage was associated with a significantly reduced odds of either moderate or severe COVID-19, these associations were prone to multiple bias, including differences in the demographic of patients presenting during wave 1 and wave 2, as well as the introduction of more standardised approaches to patient management in the second wave, including increased use of treatments such as remdesivir and dexamethasone [31-33]. Additionally, the associations were significantly abrogated by adjustment for patient demographics and overall our analysis does not support any robust relationship between the more frequently observed variants and disease severity.

This study does have limitations. Although our study contains a relatively small sample size compared to the overall reported cases in Ireland and from only one region in Ireland (the South East), it does include a significant proportion of hospitalised cases (15.2% overall) from major hospitals in a geographical location where the majority of SARS-CoV-2 cases in Ireland have been reported. As we only sequenced samples with Cq values of 30 or below, viral genetic lineages with lower viral loads theoretically may be unrepresented, although this technical limitation would apply to all analyses of WGS of SARS-CoV-2. Whole genome sequencing studies in SARS-CoV-2 are also prone to sampling bias, with greater sampling of outbreaks or to detect variants of interest. However, as the AIID cohort took an unselected approach to sampling and did not pre-screen for variants of interest, the impact of any sampling bias would have been less evident.

In conclusion, we have described disappearances of viral lineages from hospitalised COVID-19 cases during wave 1 of the pandemic in Ireland occurring alongside lockdown restrictions, and emergence of different SARS-CoV-2 lineages contributing to hospitalisations during wave 2, with phylogenetic analysis pointing to importation through travel as an important source. These findings show the utility of SARS-CoV-2 genomic epidemiology in providing insights into viral genetic lineages and variants of concern and have significant implications for reflective learning as part of pandemic control of SARS-CoV-2 infection. Whole genome sequencing of SARS-CoV-2 isolates provides critical information for the monitoring for the emergence of new viral lineages and variants of concern.

## Data Availability

Data referred to in the article are available in Table 1 & 2, Figures 1 - 4 and Supplemental figure 1.

## Acknowledgements

The authors would like to thank all the participants who provided data and samples to the AIID Cohort along with the clinical and support staff who facilitated operationally with the AIID Cohort activities. Thanks to ICHEC for providing access to their high-performance computing.

## The All Ireland Infectious Diseases Cohort Study Investigators

MMUH: Aoife Cotter, Eavan Muldoon, Gerard Sheehan, Tara McGinty, Jack Lambert, Sandra Green, Kelly Leamy. SVUH: Grace Kenny, Kathleen McCann, Neil Kelly, Matthew Blair, Rachel McCann, Claire Kenny, Cathal O’Brion, Sarmad Waqas, Stefano Savinelli, Eoin Feeney, Patrick Mallon, Pooja Varghese. CEPHR: Sarah Miles, Dana Alalwan, Riya Negi. Beaumont Hospital: Eoghan de Barra, Sam McConkey, Christine Kelly. University College Cork: Mary Horgan, Corinna Sadlier.

## The Irish Coronavirus Sequencing Consortium (ICSC)

Teagasc and APC Microbiome Ireland: Fiona Crispie, John Kenny, Paul Cotter, Calum Walsh, Elaine Lawton, Amy Fitzpatrick; Teagasc: Ewen Mullins, Michele Della Bartola, Matt McCabe; Limerick University Hospital: Patrick Stapleton, Carolyn Meaney; University College Cork, Cork University Hospital and APC Microbiome Ireland: Liam Fanning, Michael Prentice; University College Cork and APC Microbiome Ireland: John MacSharry; University College Dublin: Dr Patrick Mallon, Alejandro Leon; National Virus Reference Laboratory, University College Dublin: Gabriel Gonzalez, Michael Carr, Aditi Chaturvedi, Cillian De Gascun; NUI Galway: Simone Coughlan, Grainne McAndrew, Kate Reddington; NUI Maynooth: Fiona Walsh, David Fitzgerald; Genuity Science Ireland: Tom O’Dwyer, Lara Clarke, David Jebb, Jessica Klopp, David Kavanagh, Karl Haslam Patrick Buckley; The Royal College of Surgeons in Ireland, Dublin and Department of Clinical Microbiology, Beaumont Hospital, Dublin: Fidelma Fitzpatrick; Health Protection Surveillance Centre, Dublin and Department of Clinical Microbiology, Beaumont Hospital, Dublin: Karen Burns; Department of Clinical Microbiology, Beaumont Hospital: Jacqueline Cafferkey, Aisling Richmond, Margaret Foley; Trinity College Dublin: Jose Sanchez-Morgado; Helixworks Technologies, Ltd., Cork: Sachin Chalapati, Nimesh Pinnamaneni, Conor Crosbie, Dixita Limbachiya.

---

**Supplemental figure 1:**
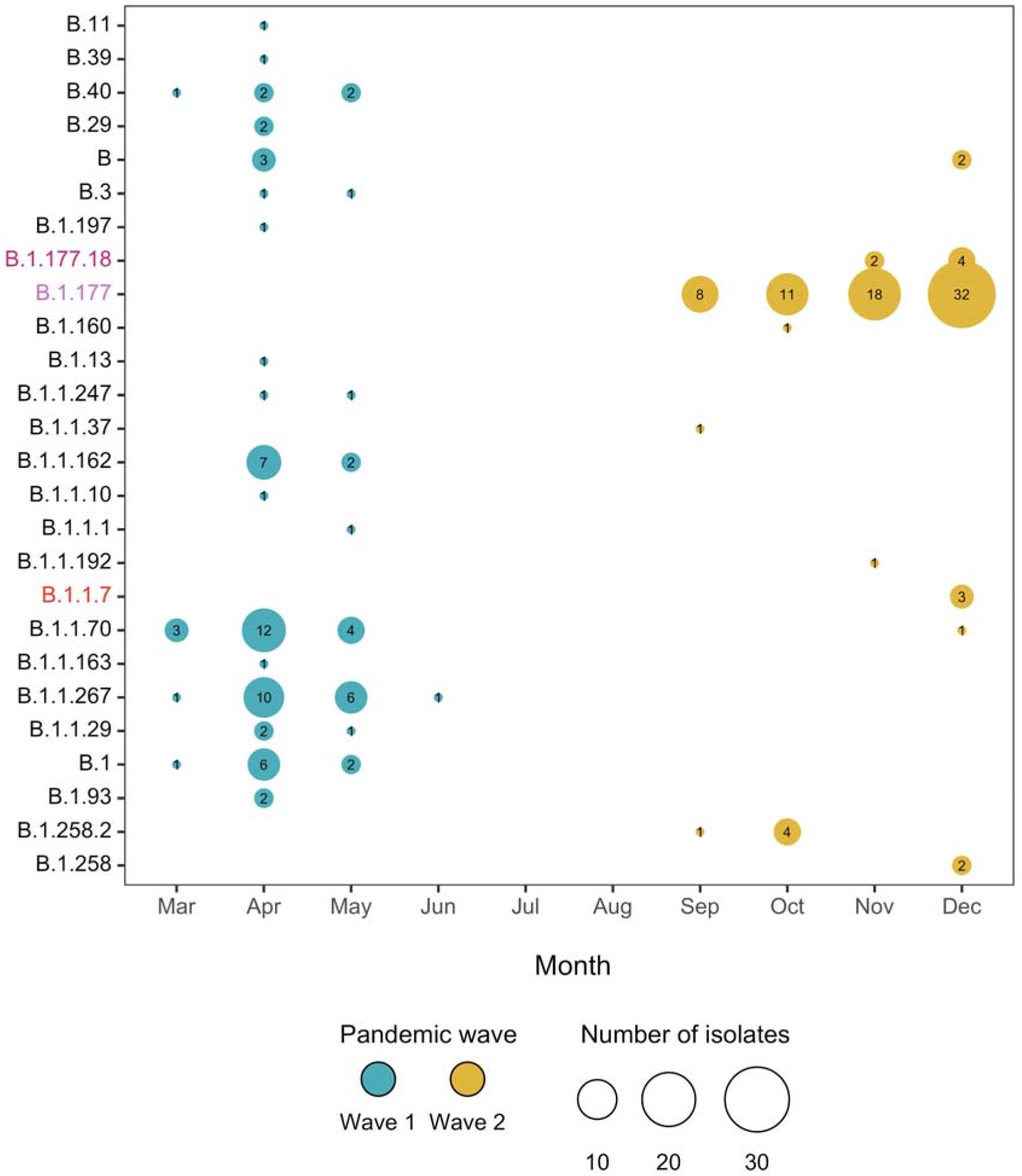
SARS-CoV-2 genome lineage classification by month between March and December 2020. Legend: The size of the circle is proportional to the number of lineages detected.

